# Longitudinal changes in motivational determinants of sustainable diets during a blended digital behavior change intervention: a mixed methods study

**DOI:** 10.1101/2025.11.28.25340360

**Authors:** Laura Bervian, Ujué Frésán, Joren Buekers, Miguel López-Moreno, Sergi Fàbregues, Claudia Teran Escobar, Guillaume Chevance

## Abstract

The adoption of sustainable diets in high-income countries is an urgent environmental and health priority. Understanding motivational determinants in behavior change interventions can contribute to clarifying the main pathways to effective dietary transformation. This mixed methods study explored longitudinal motivational determinants change during a year-long blended digital intervention aimed at promoting sustainable diet change in healthy omnivorous adults. The 22-week intervention comprised app-delivered motivational and informational text messages, as well as tailored online feedback sessions. Quantitative data were collected through repeated self-reports of diet and motivational determinants (e.g., health, environmental, ethical, price, self-efficacy, and habit strength) over 15 weeks across the year and were analyzed with linear mixed models and within-person network analysis. Qualitative data were collected through semi-structured interviews at three times: baseline, post-intervention, and six-month follow-up, and were analyzed by using framework analysis. The quantitative and qualitative findings were integrated using joint display techniques. Quantitative analyses at the group level demonstrated significant increases in environmental and ethical motivations, habit strength, and adherence to a more sustainable diet, alongside a decrease in price as motivation for food choice. No change was found in health motivation and self-efficacy. The qualitative findings provided an in-depth understanding of participants’ evolving awareness, gradual adoption of new eating habits, and varying levels of confidence in maintaining behavior changes despite contextual barriers. Individual-level results showed important heterogeneity of both motives and behaviors, and within-person network analysis showed no covariation between motivational determinant change and diet change. This study demonstrates the potential for blended digital interventions to influence motivational pathways related to sustainable diets and supports the use of mixed methods designs to track the dynamic process of dietary behavior change.

## 1. Introduction

The global food system is a major driver of health and environmental challenges (1). Current low-quality dietary patterns —characterized by the excessive consumption of calorie-dense products high in sugar, salt, and unhealthy fats, as well as animal-sourced foods, combined with insufficient intake of whole plant-based foods—contribute significantly to the increase of non-communicable diseases (NCDs), such as cardiovascular diseases, cancer, and diabetes. NCDs account for around 75% of causes of death worldwide (2,3), and are considered the most critical global health challenge of the 21^st^ century in terms of disease burden and mortality (4). Globally, in 2017, 11 million deaths and 255 million disability-adjusted life years were attributable to dietary risk factors (3). In Spain, where the present study was conducted, diet-related diseases are estimated to account for about 12% of all deaths each year, a proportion similar to that of other high-income countries (5,6).

Unhealthy diets adversely affect not only individual health but also the environment (7). The global food system plays a key role in exceeding most of the planetary boundaries, which constitute thresholds that define a safe operating space for humanity on Earth. Among the nine identified boundaries, six have already been crossed (i.e., climate change, biosphere integrity, land use, freshwater use, nitrogen and phosphorus cycles, and novel pollutants), all of which are linked to the current unsustainable food system (8). In Spain, food consumption is the main contributor to environmental impact and resource overuse, accounting for 52,1% of the consumption footprint, thereby surpassing the contributions of mobility (17,1%) and housing (16,2%) (9,10). Beyond the impact on nature itself, ecosystem disruptions have an indirect impact on human health by exacerbating food insecurity and increasing exposure to extreme weather events. This increases the risk of both infectious and non-communicable diseases, disproportionately impacting vulnerable populations (11). Notably, the environmental impact derived from the yearly food demand of Spaniards has been linked to the loss of nearly half a million healthy life years (12).

A transition toward healthy diets with low environmental impact is therefore essential and urgent (1). Such diets are predominantly whole plant-based, rich in vegetables, fruits, whole grains, legumes, nuts, and unsaturated oils, and limited in animal-based foods, especially red and processed meat, as well as products high in sugars, salt, and unhealthy fats (11,13). Beyond diet composition, environmental dietary sustainability also encompasses other behaviors, such as limiting food waste, choosing minimally packaged foods, and respecting seasonality. Additionally, the social, cultural, and economic dimensions of sustainability — by prioritizing fairly sourced products that ensure fair wages for stakeholders within the food system, and supporting small-scale economies — are essential for achieving truly sustainable diets (1,14). Broad adoption of such sustainable dietary patterns requires transformative actions at multiple levels, combining systemic structural changes through policy reforms and market incentives with effective individual-level strategies aimed at modifying eating behaviors (15–18).

Individual behavior change towards sustainable diets is complex and influenced by a multitude of factors (19–23). Key individual motivational determinants frequently explored in the literature include motivation for improving health, preserving the environment, considering fair and ethical sources, minimizing cost, deriving pleasure from eating specific foods, convenience, and sociability (15, 24–29). Beyond these broad motivations, previous studies have also shown that specific psychological constructs are significantly associated with eating behaviors (30,31). For instance, higher self-efficacy —defined as the confidence in one’s ability to perform a behavior or achieve a particular goal (32)— has been associated with healthier food choices, including greater fruit and vegetable intake (33–37), as well as with sustainable food practices, such as reducing food waste (31). Potentially relevant for sustained behavior change over time, habits —defined as cue–behavior associations learned through repetition (38) — have also been identified as a critical factor in maintaining long-term changes in eating behaviors (39).

Beyond existing observational evidence, few experimental studies have examined whether motivational determinants related to sustainable diets can be modified during interventions targeting individual behavior change, and whether a modification of these psychological determinants is associated with eating behavior changes (27,40,41). Further, most previous eating behavior interventions have mainly focused on specific food groups, such as increasing fruit and vegetable intake or reducing meat consumption and fat and energy intake, instead of adopting a global dietary approach (27,40,41). Building on a recent year-long intervention designed to promote global sustainable eating behaviors (42), this study aimed to investigate, using both qualitative and quantitative methods, longitudinal changes in psychological factors, including health-, environmental-, price-, and ethical-related motives, as well as self-efficacy and habit strength related to sustainable diets. Additionally, we investigated how potential changes in these psychological factors relate to behavior changes toward sustainable diets.

## 2. Methods

A longitudinal study using a mixed methods intervention design (43,44) was conducted between October 2022 and December 2023 to investigate the motivational factors underlying sustainable dietary behavior during a quasi-experimental, blended digital intervention. Building upon our pre-registered protocol (45) and prior research on the intervention’s feasibility and its impact on dietary outcomes (42), this paper examines the motivational aspects of behavior change. While this study is independent of previous publications, additional information on the intervention design and dietary outcomes can be found elsewhere (42,45). The reporting of the study methods and findings adhered to O’Cathain et al.’s Good Reporting of A Mixed Methods Study (GRAMMS) guidelines (46).

### 2.1 Study population

Participants were recruited through email invitations sent to employees of the Barcelona Institute for Global Health (ISGlobal), as well as through social media outreach, posters in public libraries, and word-of-mouth referrals. Eligibility criteria required participants to be fluent in Spanish, aged 18-65 years, residing in the province of Barcelona, following an omnivorous diet, and having a mobile phone that supported the study app. Exclusion criteria included being pregnant or breastfeeding, being a professional athlete, following a restrictive diet (e.g. slimming, gluten-free, or low in sugars), having a history of eating disorders, having food intolerances or allergies, having chronic illnesses that might affect eating behaviors (e.g. gastrointestinal, metabolic or endocrine conditions), not being able to make decisions about food choices independently, or already presenting eating behaviors closely aligned with the principles of sustainable diets (operationalized as a baseline score of 5 or more points on the REFRESH dietary screener (47)).

Participants received €10 per evaluation week (for a maximum of €150) as compensation for completing ≥6 out of 7 daily dietary questionnaires per week. The study protocol was approved by the Drug Research Ethics Committee (CEIm) of Parc de Salut MAR (2022/10304/I), and all participants provided electronic informed consent.

### 2.2 Study design

The intervention employed a hybrid ABA n-of-1 trial design, consisting of a 2-week baseline evaluation (A), a 22-week intervention phase (B), and a 6-month follow-up (A) (Figure 1). The intervention consisted of two components: (i) app-based text messages and (ii) individualized online feedback sessions. Quantitative data were collected over 15 evaluation weeks, during which participants self-reported their daily dietary intake via a mobile app. Qualitative data were collected through three online semi-structured interviews per participant (conducted at baseline, at the end of the intervention, and the end of the follow-up). These interviews aimed to explore motivational processes underlying behavior change, identifying perceived barriers and facilitators, and complementing the interpretation of quantitative outcomes.

**Figure 1.**
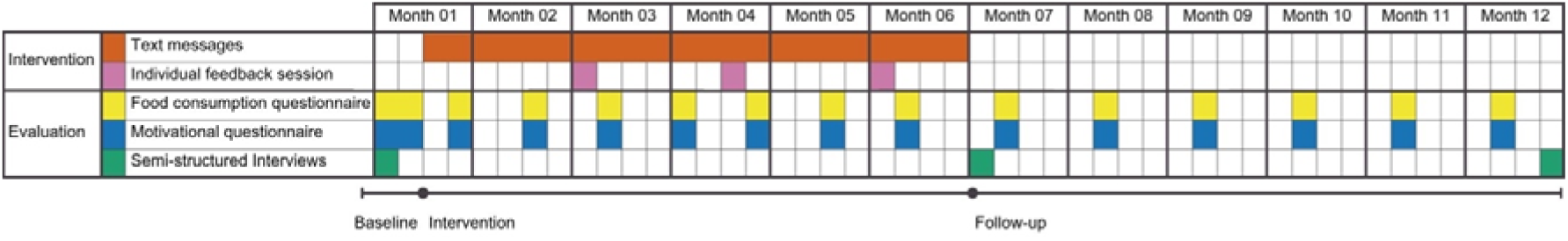
Study timeline.

### 2.3 Intervention

The intervention involved two components: generic text messages and individualized online feedback sessions. A series of 44 text messages was sent twice a week during the intervention phase using the General Data Protection Regulation-compliant app m-Path (available at https://m-path.io/landing/). The behavior change techniques (BCTs) implemented in the text messages were based on the findings from a scoping review of effective BCTs for changing eating behaviors (14) and the Compendium of Self-Enable Techniques (48). The content focused on three domains: (i) motivational messaging to encourage sustainable diets, including goal achievement tips (n = 26 messages), (ii) educational content about the health (e.g., debunking nutritional myths), environmental (e.g., reducing animal products, food waste or packaging) and socioeconomic (e.g., supporting small food system stakeholders) impacts of eating behaviors (n = 18 messages), and (iii) recipes for simple yet tasty sustainable meals (n = 4 messages). All messages underwent pre-testing with six individuals who met the predefined inclusion criteria but were not involved in the project, to ensure clarity and relevance. Refinements were made based on their feedback. Detailed information on the final message content, order, and the specific BCTs employed can be found in Additional File 1.

In addition to the text messages, participants received three individualized 15-minute online feedback sessions with a researcher (UF) specializing in dietary sustainability. These sessions provided tailored feedback based on participants’ recorded food intake from the preceding two evaluation weeks, alongside evidence-based recommendations for improving dietary sustainability. The BCTs employed during these sessions included *feedback on behavior* (i.e., based on previous measurements), tailored *instructions on how to perform the behavior to adhere to a more sustainable diet* (e.g., food substitutions, cooking tips), and *information about the health, social, environmental, and price-related aspects of dietary choices*.

No specific goals were set as part of these sessions. Although habits and self-efficacy were not specifically targeted in the current intervention, some intervention components might have influenced these two constructs, particularly self-efficacy (i.e., via behavioral monitoring and feedback (49)).

### 2.4 Quantitative measures

#### 2.4.1 Health and socio-demographic characteristics of the participants

At baseline, participants completed an online questionnaire that gathered information on gender, age, education level, household income, anthropometric measures (i.e., height and weight), and health-related factors (i.e., self-perceived physical and mental health, supplement use, and smoking status; See Additional File 2 for more information).

#### 2.4.2 Food consumption

Dietary intake was quantitatively evaluated daily over 15 weeks, with evaluations distributed across different study phases: 2 weeks at baseline, every 3 weeks during the 22-week intervention, and every 4 weeks during the 6-month follow-up period (Figure 1). As no validated brief questionnaire existed for repeated assessment of a sustainable diet (50), we developed a 10-item tool to evaluate key food groups for healthy and environmentally sustainable diets. This tool was informed by recommendations from authoritative bodies such as the EAT-Lancet Commission, the Food and Agriculture Organization, and the World Health Organization (11,16,51,52), and adapted for the Spanish context by adjusting food group examples and recommended amounts to reflect local dietary patterns and food availability. The tool generated a composite score (0-10 points, reflecting low to high adherence) to measure weekly adherence to environmentally sustainable dietary patterns. Detailed scoring criteria and rationale are provided in Additional File 3, and the Food Consumption Questionnaire used in the study is in Additional File 4.

After the intervention study, we conducted a separate validation in an independent sample of 106 adults to assess the accuracy of the questionnaire in estimating the frequency of consumption of key food groups, by comparing it with food diary records. The results indicated a small bias in food group quantification, suggesting acceptable agreement. This validation focused on the frequency estimates used to build the score, rather than the final sustainable diet score itself. The validation study is currently under revision, and the results are available at https://osf.io/st2an/.

#### 2.4.3 Motivational factors

Quantitative measures of motivational determinants included six visual analog scales (0-10), administered once during each of the 15 evaluation weeks (Table 1). These included four items measuring motives for food choices (health, environment, affordability, and ethics), adapted from eating motivation scales (26,53); one item assessing the strength of the habit related to sustainable food choices, adapted from the Self-Report Habit Index and Self-Report Behavioral Automaticity Index (54,55); and one item evaluating self-efficacy for sustainable diets (56).

**Table 1.** Items for food-related motives, habits, and self-efficacy.

| Domain | Item | Answer options |
| --- | --- | --- |
| Health | In the last 7 days, how important was the healthiness of your food when choosing what to eat? | VAS 0-10: 0=not important at all and 10=very important |
| Environmental impact | In the last 7 days, how important was the environmental impact of your food (i.e., carbon footprint, water footprint, etc.) when choosing what to eat? | VAS 0-10: 0=not important at all and 10=very important |
| Fair and ethical sources | In the last 7 days, how important were the working conditions of the producers or sellers of your food when choosing what to eat or where to buy? | VAS 0-10: 0=not important at all and 10=very important |
| Price | In the last 7 days, how important was the price of your food when choosing what to eat? | VAS 0-10: 0=not important at all and 10=very important |
| Habit strength | Selecting healthy foods with low environmental impact and from fair sources is something that I do automatically, without thinking about it. | VAS 0-10: 0=totally in disagreement and 10=totally in agreement |
| Self-efficacy | To what extent do you feel capable of following a sustainable, healthy diet? | VAS 0-10: 0=not at all confident and 10=totally confident |

Note. VAS: visual analog scale.
*2.5 Qualitative measures*

### 2.5 Qualitative measures

The qualitative assessment involved three individual semi-structured online interviews per participant, for a total of 36 interviews, conducted at baseline, at the end of the intervention, and at the end of the follow-up (see Figure 1). These three time points were selected to capture participants’ evolving experiences and motivations: the first focused on existing diet practices and intentions; the second examined perceived changes, barriers, and facilitators; and the third addressed the maintenance of behavior change. Topics discussed during the interview included typical eating patterns, food preferences, and sustainability-related behaviors such as food waste reduction, seasonal eating, and prioritizing small local markets. Interviews were conducted by the same researcher who led the feedback sessions (UF), lasted approximately 30 to 45 minutes, and were audio-recorded, anonymized, and transcribed verbatim for analysis in MAXQDA 24 (57), supplemented by additional field notes. Further details on interview content and delivery can be found in the study protocol (45), and the interview guide is available in Additional File 5.

### 2.6 Data analyses

#### 2.6.1 Quantitative

Descriptive statistics were used to analyze the characteristics of the participants. Changes in study outcomes were assessed at both the group (i.e., nomothetic analyses) and individual (i.e., idiographic analyses) levels. At the group level, changes in each outcome were assessed using linear mixed regression models with a random intercept for individuals and the week number (weeks 1 to 46) as the independent variable. Changes at the individual level were assessed using linear regressions fitted for each individual separately with a similar independent variable (the week number).

Covariations between the study outcomes were analyzed at the group-level only to maximize statistical power for that type of analysis (i.e., too few observations were available to conduct such analyses at the idiographic level). Network analyses were performed, including all study outcomes as nodes in the network: diet, motives related to prices, environmental and health features, ethical concerns, habit, and self-efficacy. We calculated the networks using partial correlation methods as described by Costantini et al. (58). In network analysis, variables (or ‘nodes’) included are connected by an ‘edge’ if they correlate after controlling for all other correlations between variables in the network. Given the limited sample size, we only computed the within-person network, capitalizing on the repeated number of observations per participant. This analysis focused on individual deviations from their average levels across all study outcomes. We computed this network by subtracting each participant’s overall average from their scores at each time point. The objective was to examine whether changes from the average level of one variable over time were associated with changes from the average level of other variables. The network analyses were performed with high specificity and low sensitivity to minimize false-positive findings.

All analyses were performed using R version 4.4.1 and R Studio version 2024.12.1. The data and code used in this study can be found on the Open Science Framework page of the project (https://osf.io/6n7s3/).

#### 2.6.2 Qualitative

For qualitative analysis, transcripts were imported into MAXQDA 24 (57) for coding and analysis. The data were analyzed using framework analysis, a structured form of thematic analysis that involves identifying key themes and organizing them in a matrix to systematically compare data across and within participants, following the steps outlined by Gale et al (59) and Spencer et al (60). In Step 1, two researchers (UF and SF) reviewed the interview transcripts to gain a comprehensive understanding of the interview data and identify topics relevant to the objectives of this qualitative phase. In Step 2, the same researchers developed an initial thematic framework based on the identified topics. This framework was organized hierarchically with subtopics within the main topics. A third researcher (MLM) then reviewed and refined this framework, using it to index the interview data in MAXQDA. Following indexing, the researcher reviewed the indexed data to ensure it was consistent with the framework categories. Step 4 involved sorting the data by content similarity, resulting in the creation of several thematic sets. In Step 5, a fourth researcher (LB) constructed a framework matrix of motivational factors, with each row representing a case (i.e., a study participant) and each column representing a subtheme. This step enabled the research team to identify thematic patterns within each case (within-case analysis) and across the cases (cross-case analysis), while preserving the context of the participants’ views. Thus, this final step allowed for a systematic abstraction and interpretation of the interview data.

#### 2.6.3 Mixed methods integration

Quantitative and qualitative results were integrated through a merging procedure using a side-by-side joint display (61,62). This strategy allowed us to compare the results from both data types for each topic examined. We assessed whether the qualitative results on motivational factors confirmed, expanded, or complemented the quantitative results. We used a joint display matrix to align qualitative themes with quantitative trajectories, identifying areas of concordance (where the quantitative and qualitative results aligned), discordance (where the quantitative and qualitative results diverged), expansion (where the qualitative results provided additional context that expanded the quantitative results), and complementarity (where motivational aspects were captured only qualitatively). An iterative, back-and-forth process was used to refine our interpretations (63), with the side-by-side joint display serving both visual and analytic purposes. This process yielded several meta-inferences synthesizing insights from the integration (64).

## 3. Results

### 3.1 Descriptive statistics

During the three-month open registration period, 94 individuals completed the eligibility survey. Of these, 12 (13%) met the inclusion criteria. Most participants (73/82, 89%) were excluded due to already adhering to an environmentally sustainable healthy diet at baseline. All 12 eligible individuals (7 men, 5 women; mean age 26 years; individual characteristics displayed in Table 2) accepted the invitation and completed the study, resulting in a 100% retention rate (flowchart is included in Additional File 6). Detailed findings regarding the intervention’s feasibility and acceptability have been reported previously (42).

**Table 2.** Individual characteristics of the participants (n=12)

| ID | 1 | 2 | 3 | 4 | 5 | 6 | 7 | 8 | 9 | 10 | 11 | 12 |
| --- | --- | --- | --- | --- | --- | --- | --- | --- | --- | --- | --- | --- |
| **Sociodemographic characteristics** |  |  |  |  |  |  |  |  |  |  |  |  |
| Gender | Male | Female | Female | Male | Male | Male | Female | Male | Male | Male | Female | Female |
| Age range (years) | 23–27 | 23–27 | 48–52 | 18–22 | 33–37 | 28–32 | 33–37 | 33–37 | 38–42 | 23–27 | 38–42 | 23–27 |
| Education | Master | Master | Doctorate | Secondary | Doctorate | University degree | Doctorate | Secondary | Master | Master | Master | Master |
| Incomes (of the family unit after taxes per month) | 1500–1999 € | 2000–2999 € | 3000–3999 € | 1500–1999 € | 2000–2999 € | 1500–1999 € | 4000 or more € | 2000–2999 € | 4000 or more € | 1000–1499 € | 1500–1999 € | 2000–2999 € |

### 3.2 Quantitative changes in food consumption and motivational determinants

Group-level results indicated a significant and positive linear weekly increase for the diet composite score [.04 (95% CI .03, .05)], environmental [.02 (95% CI .001, .03)] and ethical motives [.03 (95% CI .008, .04)], as well as habit strength .04 (95% CI .03, .06)]; all characterized by a small effect size (i.e., between .02 and .04 points increase for each evaluation weeks on a 0 to 10 scale). Price-related motives demonstrated a significant decrease over time [−0.02 (95% CI −0.04, −0.004)]. No significant changes were observed in health-related motives [.004 (95% CI −0.01, .02)] or self-efficacy [.005 (95% CI −0.008, .02)].

Individual-level trajectories varied substantially across participants. Linear regressions performed at the individual level were summarized in forest plots (Figure 2), with specific within-person changes over time shown in Figure 3. Significant increases were observed in diet score (IDs 01, 02, 04, 05, 07, 09, and 11), environmental motives (IDs 02, 09), ethical motives (IDs 05, 09), health motives (IDs 05), habit strength (IDs 02, 03, 05, 08, 09, 11), and self-efficacy (IDs 02, 05, 09). Significant decreases were observed in price motives (IDs 02, 11), health motives (ID08), and self-efficacy (ID08). These results illustrate the heterogeneity of individual responses. Additionally, for several participants, motives with non-significant results showed variability or non-linear patterns. Full model outputs are available in Additional File 7.

**Figure 2.**
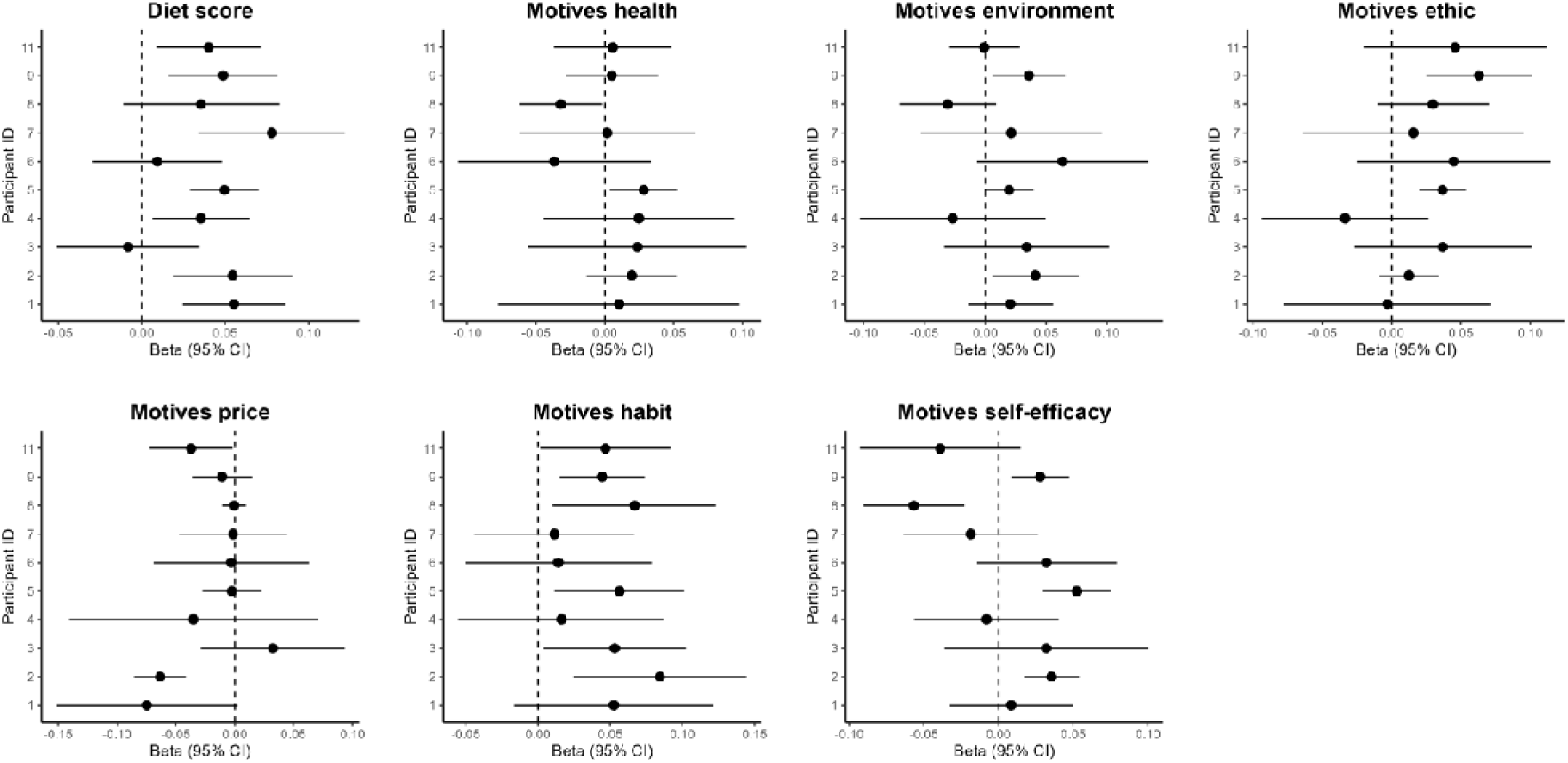
Changes for each study outcome at the individual level (progressive week is the independent variable). The x-axis shows the estimated weekly change (beta, 95% CI), and the y-axis shows participant IDs. Dots represent individual slopes, and horizontal lines indicate 95% confidence intervals.

**Figure 3.**
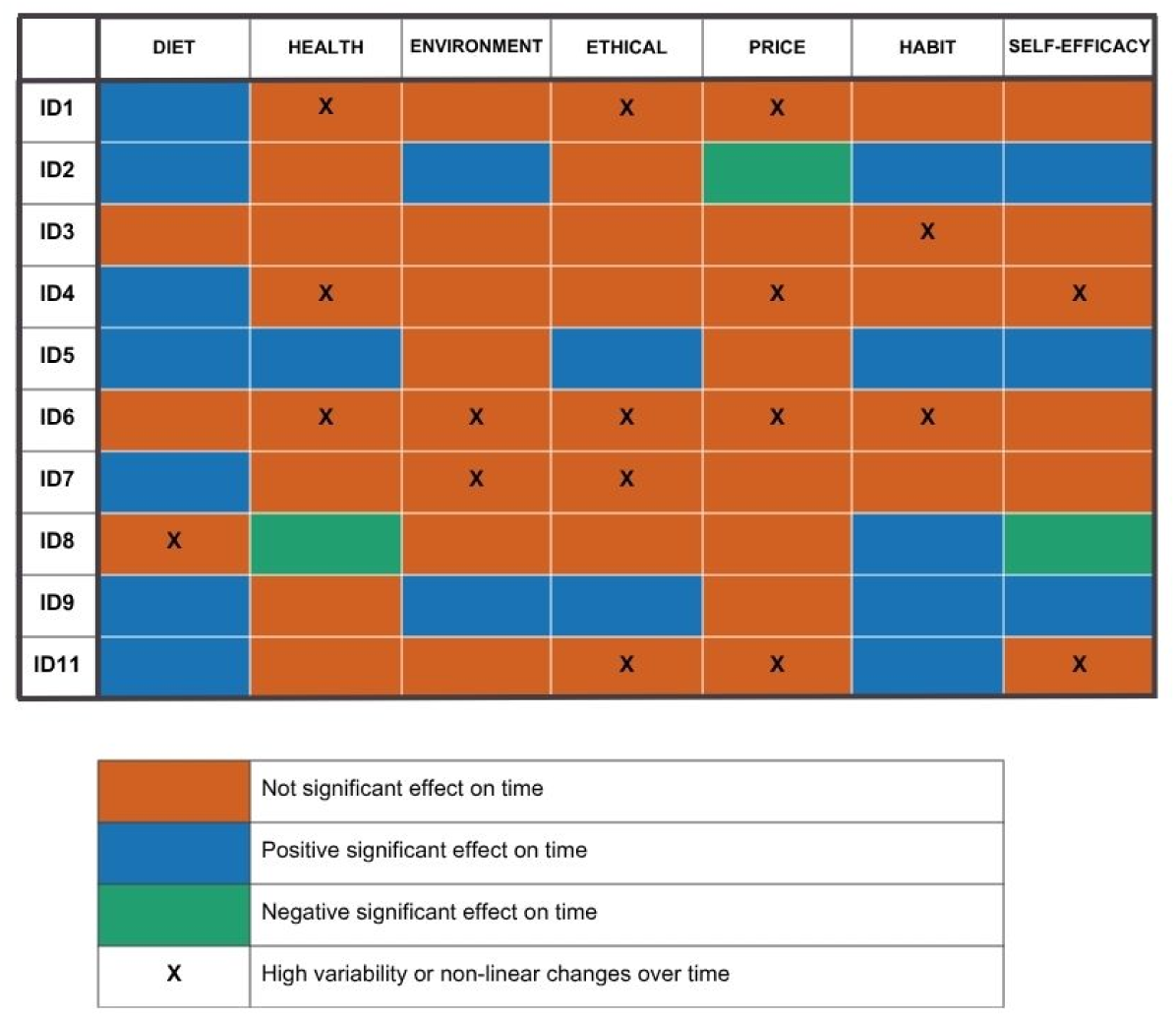
Effect of Time on Motives at the Individual-Level.

### 3.3 Quantitative associations between motivational determinants and diet composition

Figure 4 shows the results from the within-person network computed at the group-level. Positive covariations were found between all motivational outcomes, while no significant covariations were found between any motivational outcomes and diet. For example, and to illustrate, when participants reported a higher level of environmental motives at any given time point compared to their average level across all study time points, they also tended to report higher levels of ethical and health-related motives, habits, and self-efficacy at that same time point. Habits and environmental motives were the most “connected” nodes, with four significant covariations with other variables each.

**Figure 4.**
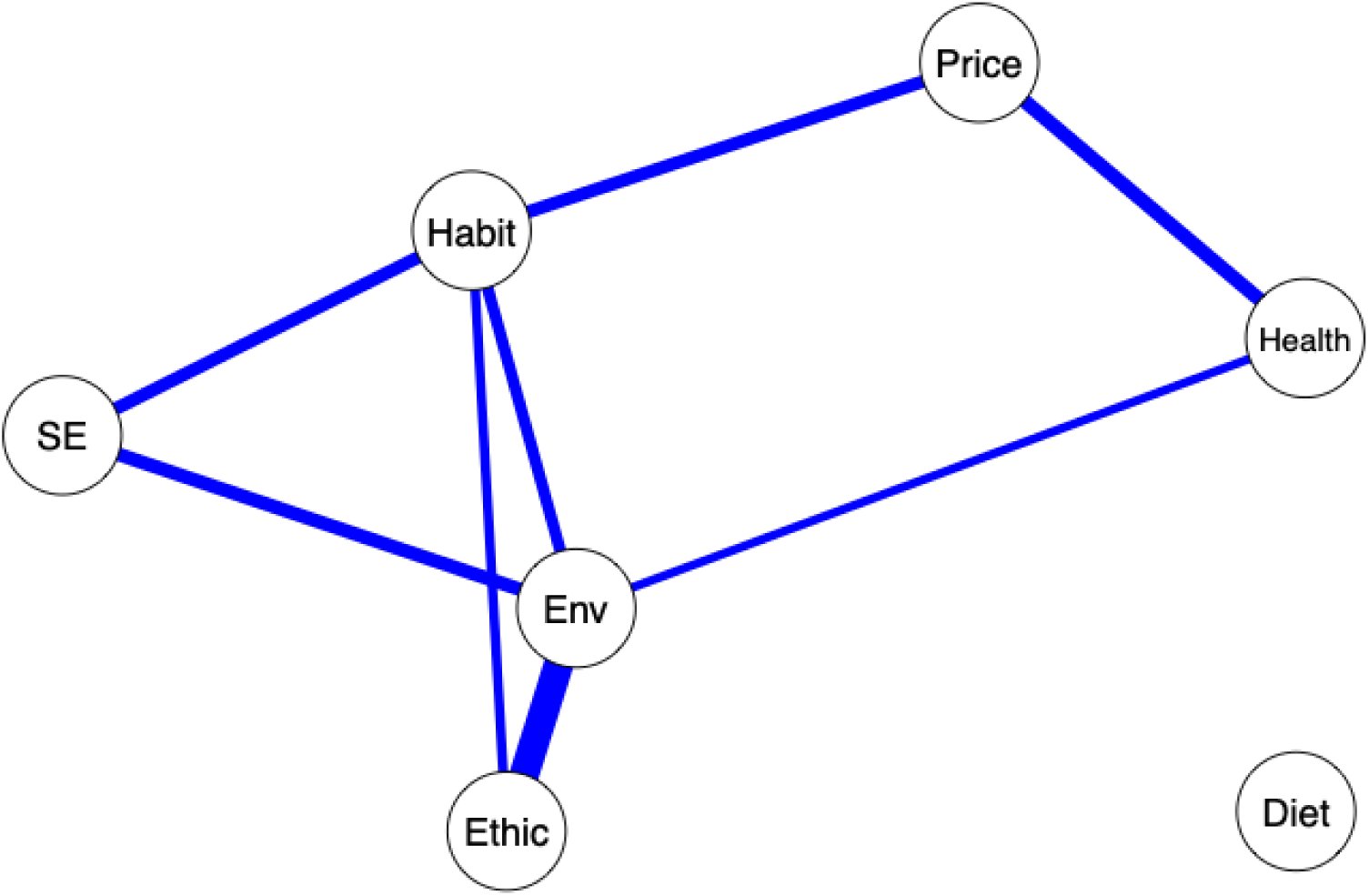
Within-person, group-level, network of the motivational determinants and sustainable diets. **Note**. SE = Self-efficacy; Env = environmental motives; blue lines indicate significant positive covariations.

### 3.4 Qualitative results

Qualitative results revealed key shifts in motivations and behaviors following the intervention. Counts (n=) indicate the number of participants who mentioned each theme during interviews. These included an increase in conscious attention to diet (n = 12), an increase in local/seasonal buying (n = 6), reduced packaging use (n = 5), and food waste reduction (n = 4). Although all participants reported an increased understanding of sustainable diets, behavioral change varied in terms of adoption and maintenance. At follow-up, participants (n = 6) reported maintaining changes, (n = 2) adopted additional changes, and (n = 5) reported partial relapse to previous habits; these categories were not mutually exclusive. Reported barriers to adoption or maintenance included social/family influences (n = 10; e.g. family preferences for meat), financial constraints (n = 7), time limitations (n = 6; e.g., long or inflexible work hours and lack of time to cook), and limited access to local markets (n = 2).

Health was cited at baseline by participants (n = 11) as one of several motives for study enrollment and dietary change, and by (n = 1) as the sole motive. Post-intervention, (n = 4) reported health benefits (e.g., improved digestion, mental well-being, higher energy levels), while (n = 2) reported negative effects (n = 1 gastrointestinal discomfort attributed to increased legumes; n = 1 increased guilt when making unsustainable choices).

Environmental motivations were another key factor for both enrollment and change, referenced by participants (n = 11) alongside health/ethics, with (n = 1) prioritizing them as their main driver, and (n = 1) dismissing their relevance in their choices. Post-intervention, (n = 3) described an increase in environmental motivation, and (n = 2) reported a “holistic shift” in their mentality and awareness of the environmental impact of food, along with a deeper relationship with nature.

Ethical motives were reported by participants (n = 3) at baseline and increased post-intervention, with (n = 5) describing increased importance of supporting local economies, (n = 4) greater attentions to labor conditions, (n = 2) to fair-trade products, and (n = 2) to animal welfare. Participants (n = 4) acknowledged ethical concerns without behavior change in that direction.

Price perceptions were mixed: at baseline, participants (n = 7) prioritized affordability, while (n = 5) valued more quality, taste, or ethics over cost. Post-intervention, (n = 4) reassessed price as secondary to sustainability, with (n = 2) recognizing that intervention strategies (e.g., seasonal eating and reduction of food waste) aided this change.

Habit strength was described at baseline by participants (n = 9) as automatic, low-awareness behaviors, with food choices often made without conscious consideration and with little attention to nutritional or sustainability aspects. Post-intervention, all (n = 12) reported increased conscious attention to diet and more routinely engaging in new sustainable behaviors such as consuming more vegetables, reducing meat consumption, or planning meals to minimize food waste. Self-monitoring (e.g., dietary and motivational questionnaires), informational messages, and personalized feedback sessions were identified as key facilitators of these changes. However, participants generally described these behaviors as not yet fully automatic, indicating early stages of routinization, with some still requiring conscious planning. At follow-up, participants (n = 6) maintained changes, (n = 2) adopted additional ones after the intervention period (e.g., reducing meat intake on weekdays or cooking simple sustainable meals at home), and (n = 5) partially returned to some prior habits, particularly regarding more effortful changes (e.g., reduced legume intake, since their family dislike and the effort of cooking two different meals; eating more cookies again because it was difficult to maintain reduction).

Regarding self-efficacy at baseline, participants (n = 6) reported an intention to change but lacked the practical means or know-how, such as specific cooking techniques (e.g., legume preparation), food substitution strategies, and meal planning skills. Post-intervention, (n = 8) mentioned the important role of applying practical tips (e.g., simple recipes, incremental change strategies), (n = 7) reported increased confidence in making dietary changes, and (n = 2) highlighted the value of the autonomy-supportive nature of the feedback sessions. All (n = 12) reported a perceived increase in knowledge related to sustainable eating and dietary change.

For detailed illustrative quotes and domain-specific results, see Table 3 and Additional File 8, with participant by subtheme coding provided in Additional File 9.

**Table 3:**
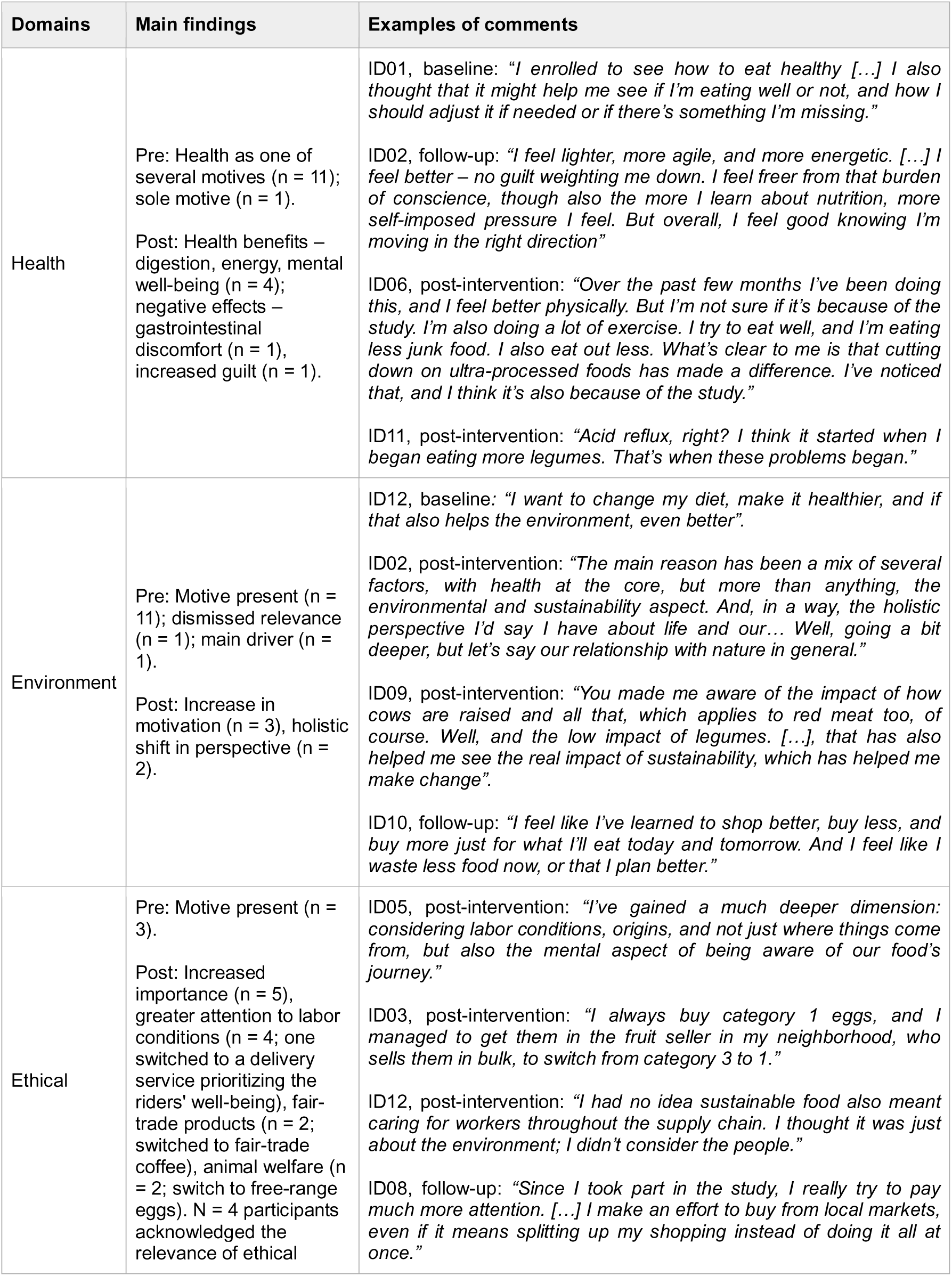
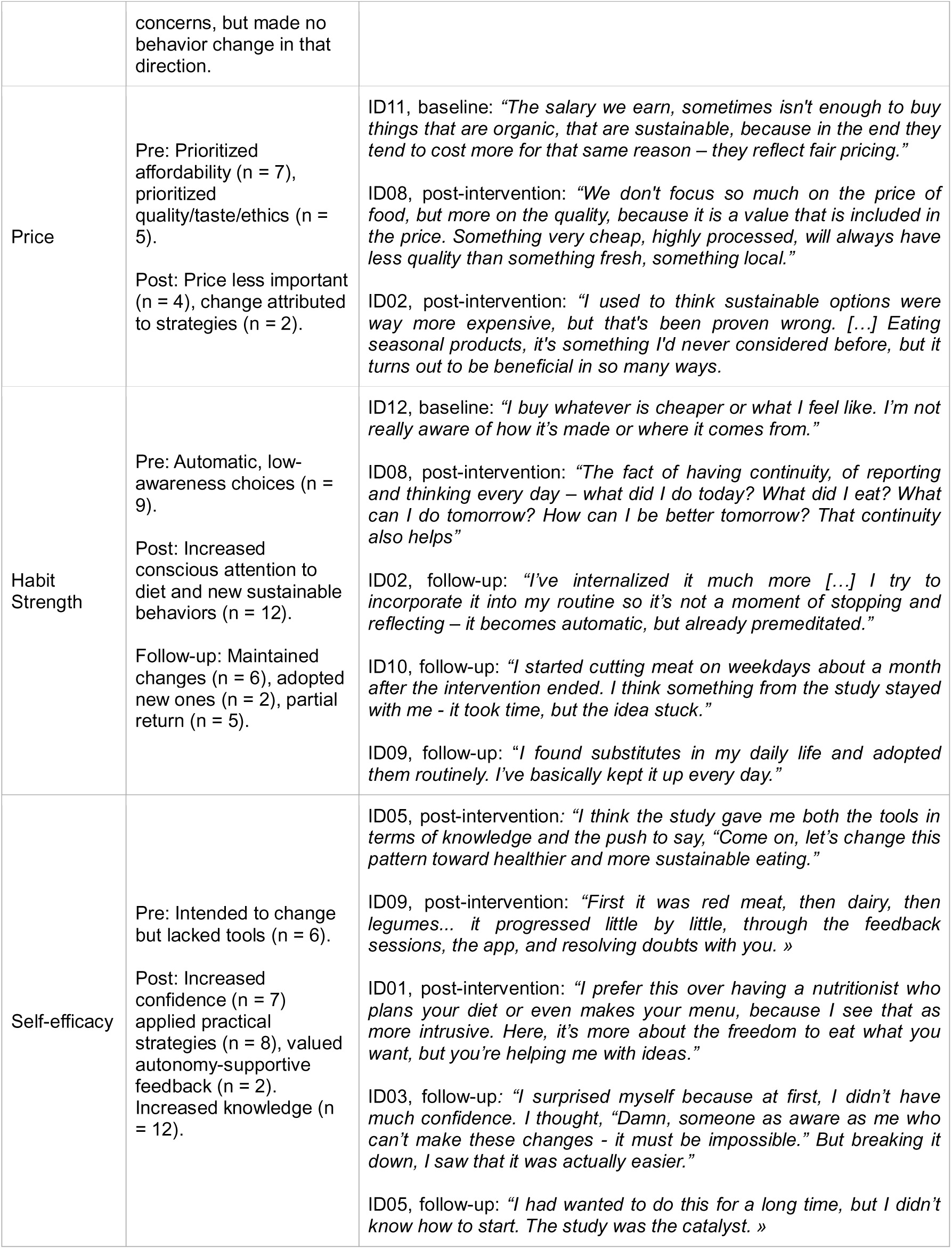
Summary of the qualitative findings for each motivational determinant. Main findings and examples of comments from each motivational determinant.

### 3.5 Mixed methods result at the group and individual levels

At the group level, quantitative and qualitative data integration revealed both convergent and divergent motivational patterns. Environmental, ethical, price-related motives, and habit strength showed concordance, with both data strands indicating strengthened motivation and engagement in sustainable behaviors (e.g., buying local and seasonal produce, reducing packaging, minimizing food waste). For health motives, qualitative findings described health as a primary and persistent driver of change, while quantitative scores showed no significant variation, indicating discordance between perceived and measured change. Self-efficacy also displayed discordance, with qualitative reports describing increases in confidence and autonomy despite no quantitative change. A side-by-side joint display at the group level is presented in Additional File 8.

At the individual level, integration revealed heterogeneous trajectories combining concordance, expansion, and discordance. Concordance was observed in participants whose quantitative improvements aligned with their qualitative reports of behavioral and motivational change. For example, participant ID02 showed increases in diet score, environmental motivation, and habit strength, while qualitatively describing a holistic shift in food choices emphasizing greater planning, seasonality and local purchasing, and the integration of new habits into daily routines.

Expansion was illustrated by ID05, who presented quantitative increases in health-and ethics-related motives, habit strength, and self-efficacy, supported by qualitative accounts of learning about health and sustainability, integrating changes into daily life, and gaining confidence in maintaining these behaviors while reflecting on broader food system issues. ID10, excluded from individual-level quantitative analysis due to insufficient data, also demonstrated expansion, with qualitative data revealing meaningful behavioral and motivational changes that developed gradually post-intervention. This participant reported adopting more sustainable practices, such as reducing meat consumption on weekdays and minimizing food waste, driven primarily by environmental and ethical concerns about worker conditions. These shifts occurred post-intervention, with the participant recognizing his need for time to process the intervention content, illustrating processes that extend beyond predefined measurement points.

Discordance was observed in some cases. For example, ID06 showed no significant changes on any quantitative measures, but reported perceived changes in eating behaviors and motivations qualitatively, such as reducing ultra-processed foods and dairy, increasing fruit and legume intake, and expressing stronger ethical concerns. These perceived changes may have been too specific or gradual to substantially influence the overall diet score or may reflect a stronger subjective perception of change than what was captured quantitatively.

Overall, individual-level integration showed three cases of concordance, six of expansion, and three of discordance, illustrating variability in motivational and behavioral change trajectories. Detailed individual level integrated results are available in Additional File 10.

## 4. Discussion

This mixed methods study examined longitudinal changes in motivational and psychological determinants over a-year-long blended digital intervention aimed at promoting sustainable diets. Quantitative results at the group level revealed significant increases in diet scores, environmental and ethical motives, and habit strength, while health motives and self-efficacy remained unchanged, and price-related motives significantly decreased. Although no covariations were found between motivational determinants and diet composition, network analysis revealed significant covariations between motivational determinants, with environmental motives and habits emerging as the most connected and central nodes. The qualitative results deepened these findings by revealing growing attention to environmental and ethical aspects of diet during the intervention, while also identifying persistent barriers regarding affordability, social influences, and time constraints.

Health motives were high at baseline and showed no significant change over time, suggesting a ceiling effect whereby participants’ health motives were maximal or near maximal at baseline, limiting potential for further improvement. The observed stability, rather than reduction, suggests that the intervention appeared to complement these pre-existing health motives without replacing them or diminishing their importance in food choices. The qualitative results confirmed health as a central reason for diet change, often alongside other motives, consistent with previous evidence indicating that health remains a primary concern in sustainable diet research and in consumers’ and health professionals’ conceptualization of what a sustainable diet is (41,65,66). Quantitative increases were observed in diet scores, environmental and ethical motives, suggesting a gradual adoption of more sustainable dietary patterns alongside a broadening of motivational drivers beyond personal health. This pattern aligns with studies showing that targeted information or reflective opportunities on the broader consequences of dietary choices can increase the salience of underrecognized motives such as animal welfare, fair labor, and ecological concerns (67–70). While health, taste and price remain primary motivators for most consumers (26), sustainability concerns can also contribute to incremental dietary changes (27,67).

Price-related motives significantly decreased over time, likely reflecting a reframing of cost as one among several decision-making criteria. Participants reported realizing that a healthy and sustainable diet could be less expensive than initially perceived, particularly when applying cost-mitigation strategies such as seasonal eating, buying in bulk, and better planning to reduce food waste – approaches promoted via messages and feedback sessions. These findings are in accordance with prior research indicating that price concerns may be reduced when sustainable behaviors demonstrate economic viability (42,71). At the same time, qualitative interviews revealed that cost remained a barrier for some participants, indicating that these concerns were not uniformly experienced. The persistence of cost-related challenges for certain individuals highlights the need for structural interventions alongside individual behavioral change efforts to further improve access to sustainable diets.

Habit strength increased significantly over time, likely due to the intervention’s high-frequency engagement (through dietary and motivational questionnaires, messages, and regular feedback), as well as its longitudinal nature, which likely contributed to habit formation, particularly through self-monitoring (38,39). Network analysis identified habit strength as a central node, covarying with environmental motives and self-efficacy. This is consistent with dual-process models where deliberative and automatic processes interact to support sustained behavior (38,55,72). The qualitative results echo these findings: participants described a shift from low-awareness routines to more intentional practices and new forms of routinization for the new behaviors. Some participants also reported an internal conflict between values and habits, leading to emotional discomfort (i.e., feeling guilty when choosing convenient processed foods despite valuing sustainability and health). This internal tension motivated changes, aligning with prior literature indicating that this mismatch can often catalyze dietary change (23).

Self-efficacy did not show significant changes in the quantitative analyses. However, qualitative interviews revealed that participants experienced increased confidence. Participants valued autonomy-supportive guidance and actionable tips, both of which are known to enhance self-efficacy and promote long-term adherence in behavioral interventions (49). This discrepancy between the quantitative and qualitative results might be attributed to the use of a single-item measure. Future studies should incorporate more comprehensive measures of self-efficacy.

Throughout the intervention, participants increasingly linked health, environmental, and ethical considerations within a multidimensional motivational framework. Rather than citing a single dominant motive, most participants described a dynamic mix of motives guiding their choices – a pattern that aligns with broader consumer research on co-occurring values such as well-being, social responsibility, and ecological concerns (73,74). Similar patterns of multidimensional motives have been described cross-sectionally (24,25) but are rarely examined longitudinally. By addressing multiple motives and behaviors simultaneously, and by emphasizing synergies (e.g., sustainable eating as both healthy and ethical), our intervention likely contributed to broadening and deepening participants’ motivational perspectives beyond what is typically captured in single-focus designs (75,76). This is further supported by the within-person network analysis, which identified significant covariations between motivational variables. However, the absence of covariations between motivational determinants and diet contrasts with the qualitative results, where participants frequently reported increases in knowledge and awareness, practical tools, and self-efficacy as important conditions in enabling dietary changes. This discrepancy may be explained by the fact that network analyses, as specified here, captured overall covariation across the full study period, whereas the qualitative interviews reflected changes occurring between baseline, post-intervention and follow-up (77,78).

### 4.1 Strengths and limitations

A strength of this study lies in the longitudinal collection of both quantitative and qualitative data, which allowed us a comprehensive understanding of the impact of the intervention on behavior change and related motivational factors. The mixed methods design added depth by integrating quantitative trajectories with qualitative insights, offering a more complete account of how changes occurred and how participants experienced them. Brief, continuous assessments implemented through the app, together with qualitative interviews conducted across three time points, provided a high-resolution and dynamic view of the changes in dietary consumption and motivation. This approach yields additional advantages, especially when compared to studies using more traditional, lower-resolution designs (e.g., cross-sectional studies or pre- and post- intervention). Our broad characterization of sustainable diets beyond the consumption of specific food groups (e.g., red meat) is also a strength.

This study has also several limitations. First, the recruitment strategy likely resulted in a relatively homogenous sample, which, together with the small sample size, limited the generalizability of the results to a broader population. Future studies should allocate more resources to recruitment strategies to include more diverse populations (e.g., individuals from lower socio-economic backgrounds). Second, to minimize participants’ burden, we used short-custom items to measure motivational determinants longitudinally. However, longer questionnaires could have provided a more detailed understanding of the changes occurring during the study. Third, the unique specificities of our design (intensive longitudinal design with a small number of participants) prevented us from testing formal mediation hypotheses, where a change in psychological constructs would explain a subsequent change in diet. Such mediations, however, should be conducted in future similar studies to identify formal mechanisms of change.

## 5. Conclusion

This mixed methods study provides novel insights into the motivational and psychological dynamics underlying dietary change during a year-long blended digital intervention. Our results indicate that, while health motives remained stable, likely due to a ceiling effect, participants increasingly integrated environmental, ethical, and cost-related considerations into their dietary choices. The qualitative findings further highlighted the role of barriers such as affordability and time constraints. Taken together, these findings highlight the complex interplay among different motivational determinants and behavior in the context of long-term sustainable dietary change.

## Acknowledgements

UF, JB and GC acknowledge support from the grant CEX2023-0001290-S funded by MCIN/AEI/ 10.13039/501100011033, and from the Generalitat de Catalunya through the CERCA Program. UF also acknowledges support from Daniel y Nina Carasso Foundation, through the Daniel Carasso postdoctoral fellowship, the Ramon y Cajal grant (RYC 2023) funded by the Spanish Ministry of Science and Innovation, and AGAUR (Generalitat de Catalunya) to the Consolidated Research Group “Sustainability in Biosystems” (no.ref. 2021 SGR 01568). JB receives funding from the grant “FJC2021-046458-I” financed by MICIU/AEI /10.13039/501100011033 and by the European Union NextGenerationEU/PRTR. Funders has no role in the conceptualization, design, data collection, analysis, decision to publish, or preparation of the manuscript.

## Ethics Approval and Consent to Participate

The research protocol of this project has been approved by the Ethics Board *Comité de Ética de la Investigación con medicamentos del Parc de Salut MAR* (number 2022/10304/I) on October 19th, 2022, and was published before starting the implementation, after having been peer-reviewed (45). All research was performed in accordance with the Declaration of Helsinki. Participants signed the informed consent form before being enrolled in the study.

## Data Availability

The datasets generated and analyzed during the current study are available in Open Science Framework: https://osf.io/6n7s3/

## Author Contribution CRediT

Conceptualization, LB, UF, GC; Methodology, UF, JB, GC; Formal analysis, UF, LB, JB, ML-M, SF, GC; Investigation, UF; Data curation, LB, UF, JB, GC; Visualization, LB, JB; Writing – original draft, LB; Writing – review & editing, all authors; Supervision, GC; Project administration, UF, GC.

## Declaration of generative AI and AI-assisted technologies in the writing process

During the preparation of this work the authors used ChatGPT in order to improve readability and language of the manuscript. After using this tool, the authors reviewed and edited the content as needed and takes full responsibility for the content of the published article.

